# A systematic review protocol of the antiviral activity of chloroquine and hydroxychloroquine against COVID-19

**DOI:** 10.1101/2020.06.13.20130245

**Authors:** Kofi Boamah Mensah, Adwoa Bemah Boamah Mensah, Varsha Bangalee, Frasia Oosthuizen

## Abstract

**Introduction:** The recent outbreak of the Severe Acute Respiratory Syndrome Coronavirus 2 (SARS-CoV-2), or COVID-19 with no approved medicines has led to global health threat. Currently, repositioning of old medicines seems the most responsible strategy for potential cure and prevention COVID-19. Hydroxychloroquine and chloroquine have shown promising efficacy against COVID-19 related pneumonia in clinical studies. However, the mode of drug action of chloroquine and hydroxychloroquine against SARS-CoV-2 infection is not clear. This review aims to gather evidence on antiviral activity and possible mechanism of drug action of chloroquine and hydroxychloroquine on SARS-CoV-2, including in-vitro, animal studies, and studies in humans.

**Method:** A structured search of five bibliographic databases namely; Medline, Web of Science, PubMed, Cochrane CENTRAL, and Google Scholar will be undertaken to retrieve studies that describe the antiviral activity and possible mechanism of drug action of chloroquine and hydroxychloroquine on SARS-CoV-2. No restrictions will be placed on publication date, but studies will be limited to only publications in English. Duplication of studies will be removed using EndNote reference manager. Three authors will screen the citations independently based on inclusion criteria. Data extraction and assessment of risk of bias will be done independently. Meta-analysis of selected studies will be done wherever suitable.

**Ethics and dissemination:** Primary data collection will not be involved in this study, hence no need for formal ethical clearance. Findings from the study will be disseminated through a peer-reviewed publication and conference meeting.

**Trial registration number:** https://doi.org/10.17605/OSF.IO/7DJMU

**Strengths and limitations of this study:**

- This study is the first systematic review to gather current evidence on the antiviral effect and mode of action of chloroquine and hydroxychloroquine on SARS-CoV-2 infection. We expect that data that will be synthesis will provide enough information to inform COVID-19 care pathways and help clinicians caring for COVID-19 patients.
- Furthermore, this systematic review will expand our knowledge on the benefits and risks of chloroquine and hydroxychloroquine in management of COVID-19 patients and identify areas of controversies, and quality assessment.
- We anticipate that there will be few studies reporting on the mechanism of drug action and antiviral effects of chloroquine and hydroxychloroquine on SARS-CoV-2 infection.

---

The recent outbreak of the Severe Acute Respiratory Syndrome Coronavirus 2 (SARS-CoV-2), or COVID-19 was initially reported in Wuhan, mainland China (Lai, Shih, Ko, Tang, & Hsueh, 2020; Wang et al., 2020). The disease has spread to other provincials in China and other countries in the world (Yao et al., 2020). In March 2020, the number of confirmed cases in Wuhan was estimated at 125 048, with 4 614 death, and >510 000 established cases in 199 countries around the world (Lythgoe et al., 2016; Rosa & Santos, 2020). The epidemic was declared by the World Health Organization (WHO) on 12^th^ March as pandemic (WHO, 2020).

Currently, it is estimated that the number of recorded cases of SARS-CoV-2 infection rises by nearly 1000 cases a day (Yao et al., 2020). With the increasing rates of morbidity and mortality, researching for prevention and cure has been a global concern (Gupta, Agrawal, & Ish, 2020). Unfortunately, no regulatory agencies have approved medicines for the treatment of COVID-19. With the current global health threat, repositioning of old medicines seems the most interesting and responsible strategy for potential cure and prevention. This is because knowledge on the medicine safety profile is known, drug interactions and posology are also well known (Colson, Rolain, & Raoult, 2020; Colson, Rolain, Lagier, Brouqui, & Raoult, 2020).

Several medicines have already undergone testing, among which hydroxychloroquine and chloroquine have shown promising efficacy against COVID-19 related pneumonia in current clinical studies (Fantini, Di Scala, Chahinian, & Yahi, 2020). Chloroquine and hydroxychloroquine have been found to prevent the in-vitro growth of SARS-CoV-2 (Wang et al., 2020). This finding is supported clinically by a study done in nearly 100 infected SARS-CoV-2 patients (Huang, 2020; Li Y, 2020). Prof. Raoult, a French virologist, has indicated that findings from a controversial non-randomized open study of 20 patients showed remarkable results on SARS-CoV-2 washout (Gautret et al., 2020; Moore, 2020).

A tweet on 21^st^ March 2020 by President Donald Trump, affirming that hydroxychloroquine “has a real possibility to be one of the biggest game-changers in the history of medicine” spark a worldwide rush for the medicine. Also, frontline healthcare workers of the pandemic are using chloroquine and its hydroxy analog under “compassionate use” or “off label” situations (Colson, Rolain, Lagier, et al., 2020; Gautret et al., 2020). However, the mode of drug action of chloroquine and hydroxychloroquine against SARS-CoV-2 infection is not clear as the medicine seems to have a wide range of likely antiviral activity (Gao, Tian, & Yang, 2020). It is therefore essential that we know the likely antiviral effect or activity and mode of drug action of chloroquine and hydroxychloroquine against the SARS-CoV-2 infection in order to be informed as we wait for findings from clinical trials and vaccine development. This review aims to gather evidence on antiviral activity and possible mechanism of drug action of chloroquine and hydroxychloroquine on SARS-CoV-2, including in-vitro, animal studies, and studies in humans.

## METHODS AND ANALYSIS

This review will follow the guidelines of the Preferred Reporting Items for Systematic Reviews and Meta-analyses (PRISMA) and conform to the standards and recommendations described by the Cochrane Collaboration. The protocol was designed based on the Preferred Reporting Items for Systematic Reviews and Meta-analyses Protocols (PRISMA-P). The study will commence in September 2020; by this time, much evidence on the use of chloroquine and hydroxychloroquine on SARS-CoV-2 would have been published. The registration of the review under Open Science Framework can be found at https://doi.org/10.17605/OSF.IO/7DJMU

### Eligibility criteria

#### Population

The systematic review will involve studies that report on participants (≥16 years) with a confirmed diagnosis of COVID-19. Randomized controlled trials (RCTs) with or without parallel design, clinical trials, case control reports and studies, cohort studies, in vitro and animal studies. Non-randomized, Quasi-randomized, and observational studies will be excluded because of the high risk of selection bias and confounding factors (Odgaard□Jensen et al., 2011).

#### Intervention

Studies that report on the administration of any dose of chloroquine or hydroxychloroquine at any frequency, any route or type of administration, and any duration of exposure or treatment. We will consider hydroxychloroquine or chloroquine in association with different interventions, if the effects of the hydroxychloroquine or chloroquine can be assessed.

#### Comparator

Included studies can have a comparator group, if any such as a placebo group, other antiviral agents’ group, other antimalaria agents’ group, other biological agents’ group, and other non-pharmacological interventions group.

#### Outcomes

The primary results of interest will be death related to COVID-19. This will be defined by the percentage of deaths from the disease to the comparison groups. Also, pneumonia-related COVID-19, which will be defined by the percentage of participants experiencing progression to pneumonia. Again, any resulting adverse effects that will be experienced by participants. Secondary outcomes will include the percentage of all-cause mortality among participants, viral clearance among participant(s) which is confirmed by polymerase chain reaction (PCR) or other approved test, percentage of participants hospitalized due to the medications, and the time-cause for participants to be symptoms-free. Pls the outcomes on the drug actions is missing.

### Search strategy

We will retrieve studies using five bibliographic databases: Medline, Web of Science, PubMed, Cochrane CENTRAL, and Google Scholar. No restrictions will be placed on publication date, but studies will be limited to only publications in English because it is the only language of communication between the authors. Further citations will be retrieved from the reference section of the included studies. Experts and authors of key publications will be contacted when necessary. Three researchers (KBM, ABBM, and VB) will independently perform the search strategy. The bibliographic software, EndNote, will be employed to organize, store, and manage all the citations and provide exhaustive and systematic search.

Firstly, controlled descriptors such as MeSH terms and their keywords were checked in the selected database. The search terms were merged using the ‘AND’ and ‘OR’ Boolean operators (Lefebvre, Manheimer, & Glanville, 2008). Then, a search strategy combining MeSH terms and keywords, such as (Antiviral OR antivirus OR anti-viral agents) AND (effect OR activity) AND (drug action OR mode of action) AND (chloroquine OR chloroquine phosphate OR hydroxychloroquine OR hydroxychloroquine sulfate) AND (Human coronavirus OR COVID-19 OR SARS-CoV-2) AND (randomized controlled trials OR randomized controlled trial OR clinical trial OR controlled clinical trial)was used.

### Study selection

The Endnote reference manager will be used to manage the selected citation by first eliminating the duplicates. This will be followed by the manual elimination of duplicates by the three independent reviewers. After the elimination of duplications, the screening of the selected studies will be done in two phases. First, screening of the titles and abstracts, and secondly, full-text screening. Inter-rater agreement will be evaluated using Cohen’s κ coefficient (McGinn, Wyer, Newman, Keitz, & Leipzig, 2004). With this statistic, values <0.4 means poor agreement, 0.4– 0.59 indicates fair agreement, 0.60–0.74 shows good agreement, and >0.75 indicate excellent agreement (Cooper, Hedges, & Valentine, 2019). The disagreement between the reviewers will be addressed through discussion or with the help of a fourth member of the team.

### Data extraction

Data will be extracted independently by two reviewers (KBM and ABBM), and any disagreement in the process will be resolved through dialogue or with the help of other members of the team. An existing proposed tool (Carlos Lopes-Júnior et al., 2016), developed using instructions presented in the Cochrane Collaboration about content and structure, will be used for the data extraction (J. Higgins, 2009). This will be done under the following areas: (1) study identification (study title; journal title; country of the study; lead author; year of publication); (2) characteristics of methodology (study design; trials method (e.g. method of generation of the random sequence, blinding methods), participant covariates e.g. (sample size, sex; age, clinical presentation at the time of enrollment, the time between diagnosis of COVID-19 and randomization, computerized tomography/magnetic resonance imaging (CT/MRI) results); study objective or hypothesis or research question; groups and controls; type of medication, length of therapy, validated measures; statistical analyses, adjustments; (3) Follow-up data (treatment allocation, date of randomization, types and doses of antivirals, biologicals, chloroquine or hydroxychloroquine,)(4) major findings or outcomes; (5) limitations and (6) conclusions. If findings in a selected article are not clear, the corresponding author will be contacted by email for further explanation or information. Microsoft Excel sheet will be used by the two reviewers to summarize the data from the selected studies independently. The excel sheets will then be checked against each other, and any disagreements will be solved by a member of the team.

### Quality assessment

The Jadad Scale will be used to assess the methodological quality of the RCTs (Jadad et al., 1996). Scores of the Jadad scale ranges from 0 to 5, with scores <3 representing a low quality and scores ≥3 representing high quality (Jadad et al., 1996). The risk of bias and internal validity of the RCTs will also be evaluated using the Cochrane Handbook for Systematic Reviews of Interventions appraisal tool V.5.1.0 (J. P. Higgins, 2008). This appraisal tool evaluates randomization sequence allocation, blinding, selective outcome reporting, allocation concealment, and completeness of outcome data. It also categorizes studies into the low, high, or unclear risk of bias. Two reviewers (KBM and ABBM) will independently evaluate and score the methodological quality of selected trials. A member of the team will settle any disagreement. The individual studies risk of bias will be presented as a narrative statement, and also as a table.

## ANALYSIS

### Descriptive analysis

The outcome of the individual studies will be present as a narrative synthesis, in which studies will be categorized and described according to purpose and design.

### Statistical analysis

This will include the following:

Antiviral activity or effects of treatment: The expected outcome from the treatment shall be the effect on death and pneumonia, which will be presented as risk ratios. A standardized mean difference (SMD) will be used for continuous data items. Also, time to clinical improvement or failure will be presented as hazard ratios. All data will be presented with 95% confidence intervals. The clinical significance of the observed effect size for each expected outcome based on determined minimal important differences (MID) will be evaluated (Langendam et al., 2013). A relative risk reduction of 5% will be determined as the MID for mortality and pneumonia outcomes. If the observed effect is presented as SMD, these will be represented as; 0.2 SMD indicating a small effect size, 0.5 SMD indicating a medium effect size, and 0.8 indicating a large effect size (Cohen, 1988). If possible, we shall convert all SMD back to their initial units to aid clinical interpretation.

We shall only conduct a meta-analysis if the participants, treatments, and underlying clinical conditions are comparable. The extent of methodological heterogeneity and clinical diversity will be determined using I^2^ statistic: I^2^=(Q−df/Q) ×100%. In the case of moderate or insignificant heterogeneity (I^2^ ≤60%), meta-analysis will be done using the random-effects model. If significant heterogeneity is observed in selected studies, the analysis will be done as a narrative synthesis examining heterogeneity through analysis of subgroups.

The effects of variation in trial characteristics: A subgroup and meta-regression analyses shall be carried out for primary outcomes of selected studies considering the following variables; hydroxychloroquine versus chloroquine, the different doses of the medicines, the treatment duration (5 days as against more than five days),

The effects of variation in participants characteristics: A subgroup and meta-regression analyses shall be performed for primary outcomes considering the following variables; age (< 60 years old versus > 60 years old), the severity of disease (severely ill versus non-severely ill) and sex.

Evaluation of reporting bias: If we identified ten or more trials in our selected study, a funnel plot and the Duval and Tweedie’s trim and fill method will be created and analyse to examine possible publication biases (Duval & Tweedie, 2000).

### Patient and public involvement

This is a systematic review protocol, and hence there will be no participant recruitment. Hence,the dissemination of findings to participants will not be relevant.

### Amendments

All amendments to this protocol will be recorded in relation to the saved searches and method analysis. This will be documented in the bibliographic databases; EndNote and Microsoft Excel templates for data collection and synthesis.

### Dissemination

The results of the systematic review will be published in an open-access journal to provide access to researchers, research groups, and academics. Also, findings will be presented at conferences, symposia, seminars, and congresses.

## Discussion

As individuals, clinicians, governments, non-governmental organizations, and different advocacy groups all over the world strive to minimize the burden of COVID-19 pandemic, they must be aware of the current scientific evidence available as the world waits for results from ongoing clinical trials. This knowledge synthesis intends to gather current evidence on the antiviral effect of chloroquine and hydroxychloroquine on SARS-CoV-2 infection. Also, the mode of action of these compounds on SARS-CoV-2. We expect that our data synthesis will provide enough information to inform COVID-19 care pathways and help clinicians caring for COVID-19 patients.

Furthermore, this systematic review will expand our knowledge on the benefits and risks of chloroquine and hydroxychloroquine in management of COVID-19 patients and identify areas of controversies, and quality assessment

Some of the strengths of this proposed study are that we have reported on the types of studies, participants, interventions, and outcomes in addition to search strategy, data sources, data extraction method (Silagy, Middleton, & Hopewell, 2002). By publishing the study protocol, we strengthen the clarity of the research approach and reduce the risk of bias, such as selective outcome reporting (J. Higgins, 2009).

Possible limitations are the heterogeneity of measures and outcomes appraised and the possibly decreased number of studies in subgroup analyses, which can impact negatively on the statistical power in the synthesis of data.

## Data Availability

All data that will be generated will be deposit at figshare and a link to the data provided.

## Disclaimer

The content is solely the responsibility of the authors

## Ethics approval

Ethical clearance is not required because the study will not involve primary data collection.

## Data sharing statement

Our protocol is a systematic review, nonetheless, we will make any unpublished data that was involved in our study available if requested.

